# Neutrophil-to-lymphocyte Ratio (NLR) as an Index of Response to Treatment in Patients with Hepatocellular Carcinoma after Receiving Trans-arterial Chemoembolization (TACE)

**DOI:** 10.1101/2023.05.16.23290061

**Authors:** Neda Shayegan, Niloofar Ayoobi, Esmaeil Mohammadi, Hajir Saberi, Faeze Salahshour, Forough Alborzi, Nahid Sadighi, Mohammad Taher

## Abstract

**Background:** Trans-arterial chemoembolization (TACE) is commonly used for patients with large, un-resectable tumors or bridge therapy in patients with hepatocellular carcinoma (HCC) prior to liver transplantation. We evaluated the response to TACE treatment in patients with hepatocellular carcinoma according to modified RECIST criteria and determined the prognostic value of neutrophil-to-lymphocyte ratio (NLR).

**Methods:** Patients with definitive diagnosis of HCC referred for TACE were studied. The response rate to TACE treatment was assessed based on dynamic MRI 28-days after treatment according to modified RECIST. The NLR value was calculated and its prognostic value was evaluated to predict the response to treatment.

**Results:** Forty patients with HCC who underwent TACE were included in the study. The response to TACE treatment in included patients was: complete response (CR) in 6 patients (15%), partial response (PR) in 16 patients (40%) and stable disease (SD) in 18 patients (45%). No progressive disease (PD) was found. Responders (CR+PR) were 22 (55%) cases. The mean NLR after treatment in the non-responder group was significantly higher than the responder group (4.2 vs 2.4, P-value=0.026). NLR values greater than 2.6 after treatment had a sensitivity of 70.6% and a specificity of 77.3% in the diagnosis of non-responders with an Area Under the Curve of 0.73 [95% confidence interval 0.58–0.89], P-value=0.011).

**Conclusion:** Compared to responders, higher levels of NLR after treatment were observed in the non-responder group. NLR level more than 2.6 after treatment is believed to be able to discriminate non-responders as a moderate prognostication tool.

## Introduction

Hepatocellular carcinoma (HCC) is among the most lethal and prevalent cancers of human body [1, 2]. Recent efforts show that the mortality rate of HCC has substantially increased in the last decades [3, 4]. Despite improvements in the treatment strategies and technology, its prognosis has remained poor as only few patients are successful candidates for radical approaches (e.g., transplantation surgery) and liver dysfunctionality [5].

Trans-arterial chemoembolization (TACE) is largely used for HCCs in their early stages. Thus, tumors of un-resectable large size tumors, or multifocal tumors without invasion of portal vein can be managed with TACE [6, 7]. Also, it is widely recommended for ‘bridge’ therapy for patients in transplantation list. Although it has improved the survival of patients, individual cases undergoing TACE represent variable prognoses [8-10]. The latter claim is believed to be relatable to notion that patients with moderate and severe conditions are greatly different based on their tumor size, markers, liver function, and general conditions. Many factors have been implemented to assess the survival and prognosis of patients under treatment with TACE, such as those related to the tumor (e.g., vascularity, portal invasion, etc.) or patient’s condition (Child-Pugh classification) [11-15].. Many scoring systems have been created for this purpose, namely ‘Assessment for Retreatment with TACE (ART)’ and ‘Alpha-fetoprotein, BCLC, Child-Pugh, and Response (ABCR)’ systems. None of the introduced systems were found to be good assessor of TACE efficacy [15-19].

In patients with HCC, inflammatory cytokines induce systemic immune response [20]. Neutrophil-to-lymphocyte ratio (NLR) has been reported to be a prognostication tool in many disorders such as ovarian cancer, colorectal cancer, HCC, and pancreatitis [21-24]. Prior effort acknowledged that higher NLR is associated with worse clinical and survival outcomes in patients undergoing TACE [25-27]. On the other hand, recent guidelines for definition of amount of response to chemical treatments have suggested utilization of the modified Response Evaluation Criteria in Solid Tumors (mRECIST) index [28]. The latter uses the density and regressing of tumor from imaging modalities. Hence, we evaluated the mRECIST and response to TACE in patients with HCC and tried to predict it with NLR.

## Methods and material

This is a retrospective cross-sectional study on HCC patients that undergone TACE therapy in Imam Khomeini Hospital Complex’s Liver Transplantation unit during 2019 and 2020. Radiologic features of the tumor were assessed from a dynamic MRI of liver one week before TACE and another one 28 days after the procedure. For some patients TACE was performed in two sessions. Measures are reported for the last session for such cases. Response to treatment was evaluated by a single expert radiologist based on mRECIST criteria. Based on the mRECIST criteria [28], complete response (CR) was referred to when no intratumoral arterial contrast enhancement in all the lesions was present. Similarly, Partial response (PR) was defined as when more than 30% reduction in sum of diameters of viable lesions from arterial phase enhancement was detectable. Sum of these two were regarded as ‘responders’ group. On the other hand, Progressive disease (PD) was classified as if more than 20% increase in sum of diameters of viable lesions from arterial phase enhancement was observable. Lastly, patients not being divisible in any of these three were considered having a stable disease (SD). Combination of these two also were called ‘non-responders’ group. Complete or partial response were regarded as ‘responders’ group and anyone else as ‘non-responder’ group. Demographic information, clinical and drug history, and laboratory reports were also collected. Neutrophil and lymphocyte counts were extracted from complete blood count with differential (CBC diff) and NLR was calculated accordingly. This study has been approved by Tehran University of Medical Sciences ethics committee (IR.TUMS.IKHC.REC.1397.245).

Qualitative variables were reported with their count and percentage. Normal distribution of quantitative values was evaluated with histograms and Shapiro-Wilk normality test. Mean and standard deviation (StD) were used for summarizing variables with normal distribution. In non-normal distribution of measures, median and inter-quartile range (IQR) was utilized. Independent samples and paired samples Student t-tests, analysis of variance (ANOVA), and Chi-square test (χ2) were performed accordingly for aim of comparison. Mean difference (MD) and standard error (SE) are used for reporting paired-samples analysis. Receiver Operator Characteristic (ROC) curve analysis and the Area Under the Curve (AUC; with 95% confidence interval (CI)) were employed for prognostication and sensitivity analysis. AUCs higher than 0.7 were regarded as good prognostic and discriminating tool. In the next step, for statistically significant ROC curves and coordinates, we used a productive function [sensitivity × specificity][29] to search for the highest yielding number and its respective NLR as the best cut-off point. Alpha level of <0.05 was interpreted significant. Statistical analyses were performed using R statistical package v4.0.3 [R Foundation for Statistical Computing, Vienna, Austria] and SPSS v24 [IBM SPSS Statistics, Armonk, NY].

## Results

In total, forty HCC were included into study. Mean (StD) age of participants was 62.3 (9.8) and 28 (70.0%) were male (Table 2). Eleven (27.5%) patients received TACE twice. Median (IQR) of tumor size was 39.5 (29.7) millimeters with positive skewness before TACE. Most patients were bicytopenic or pancytopenic before TACE based on laboratory results. White blood cells (WBCs) changes before and after TACE were all insignificant (Table 3). Mean NLR was 2.3 (1.0) before TACE and 3.2 (2.6) after TACE, representing a significant decreasing pattern (MD [SE] = -0.9 [0.4], P-value = 0.036).

**Table 1.**

| **Table 1** | Before TACE | After TACE | P-value |
| --- | --- | --- | --- |
| Age (mean (SD)) | 62.3 (9.8) | - |  |
| Gender (Males count (percent)) | 28 (70.0%) | - |  |
| Lab data |  |  |  |
| WBC | 3671.5 (521.1) | 3708 (472.3) | 0.698 |
| Neutrophil | 2468.1 (480.2) | 2537.6 (486.5) | 0.438 |
| Lymphocyte | 1203.4 (356.7) | 1099.1 (480.7) | 0.267 |
| NLR | 2.3 (1.0) | 3.2 (2.6) | **0.036** |

**Table 2.**

| **Table 2** | Responders | Non-responders | P-value |
| --- | --- | --- | --- |
| Age (mean (SD)) | 57.0 (6.5) | 68.7 (9.3) | **<0.001** |
| Gender (Males count (percent)) | 16 (72.7) | 23 (66.7) | 0.738 |
| Lab data before TACE |  |  |  |
| WBC (mean (SD)) | 3680.6 (436.5) | 3685.4 (619.0) | 0.977 |
| Neutrophil | 2508.2 (453.6) | 2428.6 (509.2) | 0.604 |
| Lymphocyte | 1172.3 (273.7) | 1256.8 (438.3) | 0.604 |
| Hemoglobin | 10.4 (1.3) | 10.4 (1.3) | 0.901 |
| Platelet (10<sup>3</sup>) | 95.5 (9.5) | 99.8 (9.8) | 0.173 |
| NLR | 2.3 (0.9) | 2.2 (1.2) | 0.792 |
| Lab data before TACE |  |  |  |
| WBC (mean (SD)) | 3731.4 (433.2) | 3679.4 (531.0) | 0.738 |
| Neutrophil | 2396.1 (470.2) | 2720.6 (457.1) | **0.037** |
| Lymphocyte | 1269.3 (442.2) | 878.8 (448.2) | **0.010** |
| NLR | 2.4 (2.0) | 4.2 (2.9) | **0.026** |

Considering mRECIST criteria, 6 patients revealed a CR, 16 patients PR, SD in 18 patients, and no one showed to have PD. As a result, 22 (55%) of patients were responders. Mean age of non-responder patients (68.7 [9.3]) was significantly higher than responders (57.0 [6.5], P-value < 0.001). Lymphocyte counts were significantly different between responders and non-responders after TACE; i.e., in non-responders group it was 878.8 (448.2) which was lower than responders (1269.3 [442.2], P-value = 0.010) group. In contrast, neutrophil counts of non-responders (2720.6 [457.1]) was higher than their counterparts (2396.1 [470.2], P-value = 0.037). Post-TACE NLR calculation revealed an almost ×2 difference, with 4.2 (2.9) for non-responders and 2.4 (2.0) for responders (P-value = 0.026). Such variations were not observed in pre-TACE and other measurements (Table 4). ROC curve analysis revealed that NLR calculation before TACE procedure has no discriminating and prognosis ability (AUC = 0.38 [95% CI 0.20 – 0.57], P-value = 0.221) opposed to post-TACE calculation (AUC = 0.73 [95% CI 0.58 – 0.89], P-value = 0.011). The mentioned opinion indicates that higher NLR measurements after TACE are related to non-response based on mRECIST categories. Post-hoc analysis of significant ROC curve discloses a cut-off of 2.6 with 70.6% sensitivity and 77.3% specificity.

## Discussion

The main finding of current study is that NLR calculation 28-days after TACE is a moderate prognosticator of poor response to treatment and a NLR value of 2.6 is the optimal value for dichotomization of good-poor outcomes with 70.6% sensitivity and 77.3% specificity. No other cell count showed such discriminatory power. Pre-TACE analysis of NLR also was not useful.

HCC rate has been increasing in recent years as populations are ageing and chronic disorders of liver are getting more prevalent and communities are transitioning from communicable diseases toward non-communicable disorders [30-32]. Furthermore, advancement of technology and diagnostic tools have led to diagnosis of cancers in earlier stages. Although the latter has corresponded to early detection of hepatic masses, there are still remarkable portion of tumors that present with encasement and invasion of portal vein and other structures [23]. Total resection of tumor is not feasible in these group of cases and application of adjuvants to reduce the tumor size and invasion amount are considered before surgical intervention. TACE, radiofrequency ablation (RFA), and microwave ablation (MWA) are newly introduced to achieve aforesaid goal [33].

New technologies have improved the quality of care of cancer patients in recent decades [34, 35]. TACE is one of the commonest commodities to treat patients with large un-resectable HCC tumors. Also this method is widely used for bridge therapy of HCC in transplant candidates. Although, it does not completely eradicate tumor and the rate of recurrence is high. As a results, adjuvant therapy with other modalities is primarily taken [36]. In a large systematic review and meta-analysis on 12372 patient it has been revealed a 52.5% successful rate of treatment for TACE. Additionally, mean survival duration of patients was 19.4 months and five-year survival of 32.4%, represent its effectiveness [37]. Is our sample of cases, it is observed that in total 55% of patients have benefited (responded) from TACE, a proportion similar to previous studies. Many prognostication models and tools have been designed to assess the response to treatment among TACE receiving patients. The mRECIST criteria is a valid set that comprehensively investigates the amount of necrosis in tumor by dynamic magnetic images [28]. In a study on 245 patients receiving TACE treatment, it was identified that survival rate is much worse in non-responders (SD + PD) [38]. In a similar effort, it was claimed that mRECIST scores are correlated with survival [39].

We found the NLR is correlated with mRECIST categories and can be used as an adjunct to it. Other studies have reported controversial findings. One survey has mentioned that pre-TACE NLR is associated with treatment response while in our sample this was not detectable [40, 41]. Furthermore, many studies have shown the association of NLR and prognosis of patients [27, 42]. Increased lymphocytes is an antitumoral response to malignant cells. But, higher neutrophil count, and subsequently increased NLR, is a proxy of suppressed immune system which permits angiogenesis and invasion of tumoral cells [43-45]. In a study on NLR’s dynamic change after TACE, it was shown that NLR had increasing trend for the first three months and then significantly decrease to lower values [46]. All together, the controversial findings might be related to the timing of taken samples and a dynamic assessment of NLR can be more informative [46, 47].

## Limitations

Being limited to one center’s patients and referral nature of our center may have influenced our results and homogeneity of sample. Moreover, short follow-up and cross-sectional assessment of only one NLR calculation were among other limitations of this work. Low sample size was also a major drawback and withheld us from performing complementary analysis and modeling to adjust for confounders and evaluate interactions.

## Conclusion

In conclusion, TACE has become a fundamental part of HCC treatment nowadays. Efforts should be made in clinical and scientific societies to understand the tumoral behavior and associated factors to invasive approaches in priori to reduce undesired outcomes and selection of best-responding patients during pre-operative sessions. Combination of neutrophil and lymphocyte count can be an adjunct to mRECIST in post-TACE procedure. Baseline NLR and follow-up crude count of blood cells have not shown promising results. Although NLR can be used in selected cases, its trend and serial measurement are more informative. Larger studies with follow-up data and adjustment for confounding contributors may have more accurate results with predictive features.

## Data Availability

All the material and data used in this manuscript can be provided through official request to the corresponding author.

## Ethics statement

This study has been approved by Tehran University of Medical Sciences ethics committee (IR.TUMS.IKHC.REC.1397.245) and all participants were consented for participation.

## Authors’ contribution

NS performed the study and prepared the manuscript. NA, FS, NS, and HS performed TACE and participants recruitment, interpreted the results, and approved the final manuscript. EM performed statistical analysis, prepared the manuscript, and approved the final version of manuscript. FA interpreted the results and approved the final version of manuscript. MT conceptualized the study, supervised it, approved the final version, and is the corresponding author of this work.

## Acknowledgement

Authors would like to thank the participants and their families as well as all staff and other researchers of this work.

## Appendix

Not applicable

